# Analysis and Interpretation of Somatic NMD-Escaping Variants in Oncogenes and Dual-Function Genes across Adult and Pediatric Cancer Cohorts

**DOI:** 10.1101/2025.11.20.25340490

**Authors:** Mohammad K. Eldomery, Karissa M. Dieseldorff Jones, Maria Namwanje, Runjun D. Kumar, Jiaming Li, Mark R. Wilkinson, Lu Wang, Jeffery M. Klco, Li Tang, Jennifer L. Neary, Sharon E. Plon, Patrick R. Blackburn

## Abstract

Interpreting protein-truncating variants predicted to escape nonsense-mediated decay (NMDe) based on the 50-bp rule is challenging due to their variable consequences. The Clinical Genome Resource, Cancer Genomics Consortium, and Variant Interpretation for Cancer Consortium recommendations focus on tumor suppressor genes where NMDe variants may result in loss-of-function. However, guidance for interpreting NMDe variants in oncogenes and dual-function genes remains limited. To address this gap, we screened the Catalogue of Somatic Mutations in Cancer, focusing on oncogenes and dual-function genes with ≥10 NMDe variants supported by published functional studies. This analysis prioritized 15 genes exhibiting two distinct NMDe patterns resulting in gene activation. The first pattern and gene examples involve NMDe variants causing loss of C-terminal regulatory regions that mediate protein inhibition and/or degradation, frequently manifesting as attenuated cell surface receptor internalization/degradation (e.g., *CSF3R*) or increased intracellular protein stability (e.g., *CCND3*). The second is driven exclusively by frameshift NMDe variants that generate novel peptide fragments that alter protein interactions (e.g., *CALR*). Interrogation of these genes in the St. Jude Children’s Research Hospital clinical genomics pediatric cohort identified 119 NMDe variants across 8 genes in ∼3% (113/3,492) of unique patient samples, with 118 of these classified as likely oncogenic or higher (99 relating to the first pattern and 19 to the second). One variant was classified as “uncertain significance”. Our data emphasize the need to integrate gene function, variant type, and effects on the C-terminus to comprehensively evaluate somatic NMDe variants and predict their consequences across adult and pediatric cancer cohorts.

## Introduction

Nonsense-mediated decay (NMD) is a translation-dependent cellular process that targets and degrades mRNAs containing premature termination codons (PTCs) introduced by protein-truncating variants (i.e., PTC-variants).^1^ NMD prevents the synthesis of potentially harmful proteins that might have dominant-negative or gain-of-function effects, and is the mechanism by which PTC-variants may cause loss-of-function (LoF).^1–3^ PTC-variants include genetic alterations such as frameshifts, nonsense, intragenic deletion/duplication, or splice alteration with subsequent change to the open reading frame.^4^ The position of the PTC within the mRNA may determine whether or not the mRNA will escape NMD, commonly known as the ‘50-bp rule’.^5,6^ Briefly, PTCs more than 50-55 nucleotides upstream of the last exon-exon junction activate NMD, leading to mRNA degradation. Conversely, mRNAs harboring PTCs less than 50-55 nucleotides from the last exon-exon junction, or within the last exon, escape NMD and may produce a protein with an altered or truncated C-terminal domain.

Interpreting which PTC-variants are NMD-subjected (NMD-triggering) is generally more straightforward in the germline context, where LoF is a known disease mechanism, or in the somatic context, as in the case of tumor suppressor genes (TSGs).^7–9^ However, the consequences of NMD-escaping (NMDe) variants are challenging to predict in the absence of functional studies, because the functional importance of the truncated C-terminal region will vary by protein.^10,11^

In well-established genes where LoF is the disease mechanism, the Clinical Genome Resource (ClinGen) outlined a comprehensive framework to enhance curation of germline NMDe variants according to the 50-bp rule.^12^ According to the ClinGen framework, NMDe variants are interrogated to determine their consequences and then assigned the appropriate evidence strength based on whether they alter a critical region of the protein that may cause LoF. In the absence of a known functional role for the C-terminus, genes are examined to determine exon enrichment for high-frequency LoF variants using population data, such as the Genome Aggregation Database (gnomAD), the biological relevance of gene transcripts impacted, and the percentage change in protein length. Subsequent joint recommendations by ClinGen, the Cancer Genomics Consortium (CGC), and the Variant Interpretation for Cancer Consortium (VICC) further refined the ClinGen framework with regard to somatic NMDe variants in TSGs.^9^ However, despite the prevalence of NMDe variants that may activate oncogenes and genes demonstrated to have dual functions depending on the tumor context (here referred to as dual-function genes), guidance for curating these variants remains limited in both the joint recommendations as well as the Association for Molecular Pathology, American Society of Clinical Oncology, and College of American Pathologists guidelines^8,13–15^

Building on the ClinGen/CGC/VICC joint recommendations, we screened the Catalogue of Somatic Mutations in Cancer (COSMIC) to flag well-established cancer genes harboring ≥ 10 somatic NMDe variants based on the 50-bp rule.^16^ To characterize patterns of NMDe variants that may cause activation of cancer genes, the analysis focused on genes functioning as oncogenes and dual-function genes for which functional data from prior studies are available. We subsequently utilized data from the St. Jude Children’s Research Hospital (SJCRH) clinical genomics pediatric cohort as an independent dataset and evaluated the implications of these findings on the classification of somatic NMDe variants.

## Materials and Methods

### COSMIC Dataset Annotation and Curation

The variant call format (VCF) file from COSMIC (Complete Targeted Screens Mutant, version 100, downloaded on 06/08/2024) was annotated using the Ensemble Variant Effect Predictor (VEP, version 111) and GRCh38.^17^ Our analysis focused on nonsense and frameshift PTC-variants occurring within Matched Annotation from NCBI and EMBL-EBI (MANE) select transcripts as a surrogate for biologically relevant transcripts.^18^ Following the Human Genome Variation Society (HGVS) guidelines, the nonsense variants noted here refer not only to single-nucleotide substitutions but also to deletions, insertions, duplications, or insertion-deletions (indels) at the DNA level that result in an immediate stop codon.

We performed an extensive manual curation of COSMIC data to remove data from cell lines and samples with duplicate variants. In addition, samples annotated in COSMIC to high microsatellite instability or studies harboring samples characterized by elevated tumor mutation burden/hypermutation—such as the MSK-IMPACT cohort described by Zehir and colleagues — were excluded to reduce potential overrepresentation of passenger truncating variants.^19^ We also excluded samples obtained from the same individual at different anatomical sites in clonality studies.^20^ Across several iterations, merged phenotypes were reviewed to remove variants in cancer genes without a defined or well-established genotype–phenotype relationship (e.g., carcinomas involving *CALR*). For a list of all variants filtered out, refer to Table S1.

### NMD Prediction in COSMIC

NMD status based on the 50-bp rule for PTC-variants was predicted using two existing tools: the VEP NMD plugin and the aenmd R package.^21^ Additionally, a custom script was developed to identify NMDe variants according to the canonical 50-bp rule. This script utilized Ensembl BioMart data to retrieve exon location information and the position of the last amino acid in the penultimate exon or to determine whether the variant lies within the last exon. Based on these inputs, the custom script determined the NMD status of PTC-variants (NMD-triggering vs. NMD-e) relative to the 50-bp rule. Variants were classified as NMDe if at least two of the three tools were concordant. Variants predicted by the VEP NMD plugin or aeNMD algorithms to escape NMD based on non-canonical rules (e.g., located within 150 bp from the initiation codon) were excluded from this analysis. The Ensembl VEP Downstream plugin was used to estimate how a frameshift variant alters the resulting protein sequence of a transcript.

### Gene and Variant Manual Curation

Previous statistical frameworks for evaluating NMDe in cancer genes excluded frameshift variants because accurate estimation of background mutation rates for indels remains challenging.^5,22^ Therefore, we adapted a practical and clinically oriented strategy by focusing on genes harboring a minimum of 10 NMDe variants that have published gene- and variant-level functional studies. Manual curation began with defining each gene’s role in cancer and the underlying mechanism of action. Genes were annotated using the COSMIC Cancer Gene Census (CGC, v101) and OncoKB (updated on February 17^th^, 2025), categorizing them as TSGs, oncogenes, or dual-function genes.^23,24^ Genes with discordant designations between COSMIC CGC and OncoKB were manually reviewed and curated. Subsequently, we analyzed variant types and evaluated their effects on the C-terminus based on functional studies, correlating findings with known or associated tumor phenotypes. ProteinPaint, an interactive web application, was used to visually inspect the location of the variants with respect to the protein.^25^ All interactive ProteinPaint-generated HTML files are available (https://github.com/meldomer). Genes were interrogated using the gnomAD (v4.1.0) to evaluate exonic enrichment for high-frequency germline PTC-variants.

### Verification using SJCRH Clinical Genomics Cohort

The ClinGen/CGC/VICC recommendations focus on the interpretation of PTC-variants, including NMDe variants, in TSGs. The SJCRH clinical genomics cohort was used as an independent validation set to confirm the patterns of NMDe variants associated with gene activation identified in the COSMIC analysis and to assess the classification of NMDe events in oncogenes and dual-function genes. This cohort differs from COSMIC in that it predominantly includes tumors from pediatric patients. We queried the SJCRH cohort for genes prioritized within the COSMIC analysis. Sequencing was performed as previously described.^26,27^ NMD status was determined as outlined above, followed by manual review

We assessed the applicability of evidence codes such as OVS1_Very Strong (8 points) and OP4_Supporting (1 point) in these gene categories.^9^ The OP3_Supporting is typically applied for missense variants occurring in a hotspot database (https://www.cancerhotspots.org) with fewer than 10 observations. The ClinGen/CGC/VICC recommendations advised caution for truncating variants, and if the variant is underrepresented, suggested the usage of COSMIC or tumor-type–specific study. As such, we evaluated the feasibility of applying OP3_Supporting (1 point) when NMDe variants were detected in a tumor type where the gene is known to be associated with oncogenesis.

## Results

### Annotated and Curated COSMIC Dataset

Annotation of the COSMIC database revealed a total of 87,593 nonsense and frameshift PTC-variants in 55,282 unique samples across 548 unique genes/transcripts (Figure 1). After COSMIC data curation as outlined above and through several iterations of sample and phenotype review, a total of 26,142 PTC-variants from 11,761 samples were removed (Table S1). The remaining 61,451 PTC-variants were identified across 44,630 unique samples in 512 genes. Demonstrating the relevance of this variant type, we found that ∼36% (22,174/61,451) were predicted to be NMDe, occurring in 19,942 unique samples and spanning 279 unique genes/transcripts (Figure S1; Table S2). The remaining ∼64% (39,277/61,451) were predicted to undergo NMD in 28,356 unique samples. Analysis of genes with a minimum of 10 NMDe variants prioritized 84 genes accounting for 97% (21,577/22,174) of NMDe variants and detected in 19,444 unique samples.

**Figure 1.**
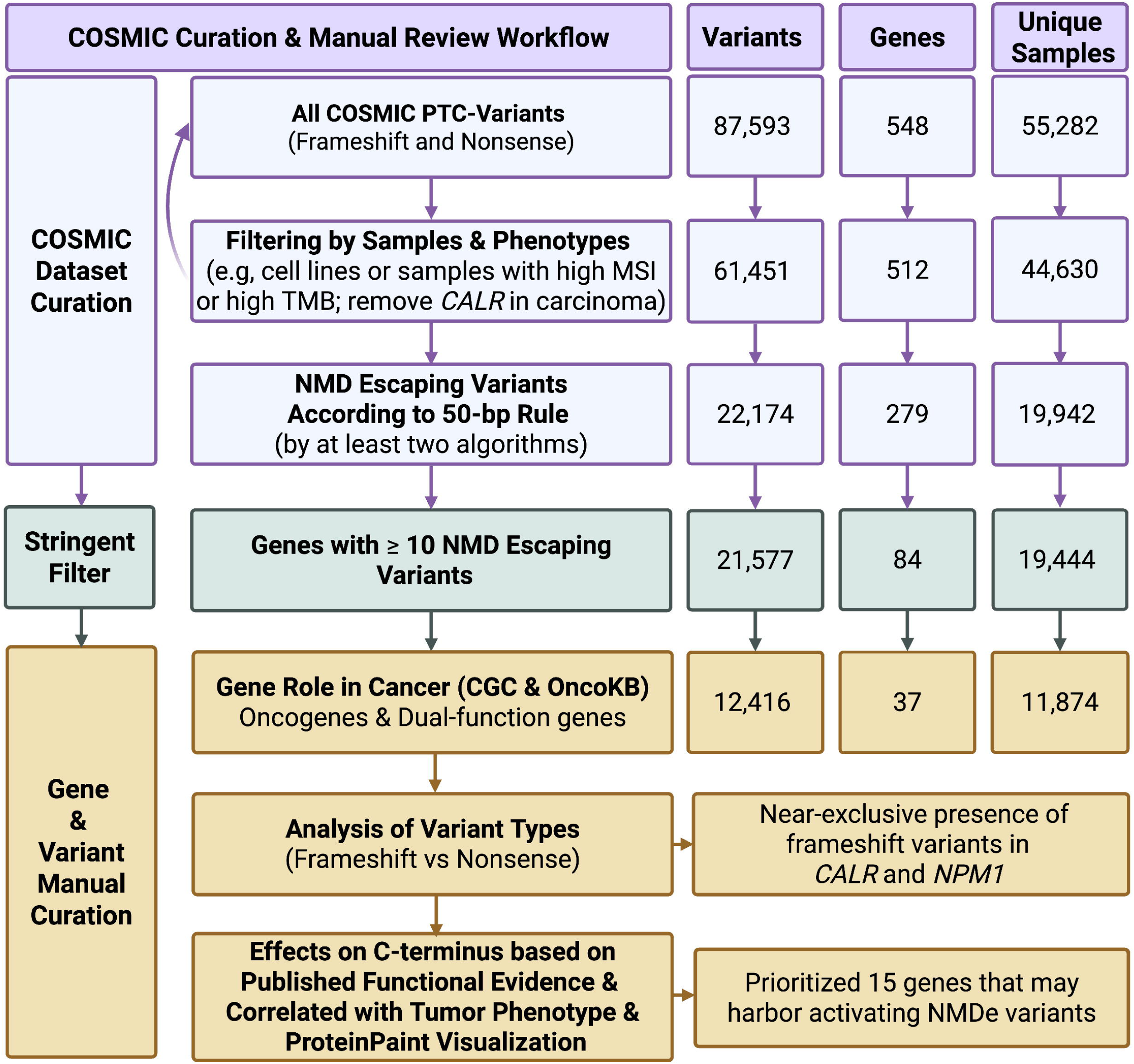
Workflow for COSMIC curation, gene and variant filtering. COSMIC annotation identified 87,593 nonsense and frameshift PTC variants across 548 genes. Curation included iterative sample and phenotype review (curved arrow). Genes with ≥10 NMDe variants (n=84) captured 97% of all NMDe variants. Manual review included curating the gene function, identifying 37 oncogenes and dual-function genes with NMDe variants and evaluating variant types, C-terminal effects and visualization by ProteinPaint.

### Curating the Gene Role in Cancer

Among the 84 evaluated genes, gene function annotations were concordant between CGC and OncoKB for 63 genes, discordant for 14 genes and unavailable in OncoKB for 7 genes (Figure 2; Table S3; Table S4). *NPM1* was annotated as a TSG in OncoKB but as an oncogene in CGC. Given that the oncogenic role of *NPM1* in acute myeloid leukemia (AML) has been attributed to both tumor suppressor and oncogenic activities, we designated it as a gene with dual roles (see sections below).^28–30^ *RUNX1*, annotated as a TSG in OncoKB and as a dual-function in CGC, has been previously reported to possess context-dependent dual roles; thus, we also categorized it as a dual-function.^31,32^ Finally, four genes (*BCORL1, CREBBP, RAD21* and *TP53*) with discordant annotations were designated as TSGs based on the majority of the evidence.^33–36^ *ASXL1* and *BCL10* genes were both annotated as TSG in OncoKB and CGC. The native, full-length BCL10 protein is known for its pro-apoptotic role.^37^ However, given its role in the activation of NF-kB signaling driven by proximity effect/structural variation or by protein truncations in mature B cell lymphomas (sections below), the final designation was a dual-function gene.^38–40^ Initial studies showed that knockout of *ASXL1* in mice results in features of myelodysplastic syndrome and therefore was regarded as a tumor suppressor gene. However, additional research studies suggested *ASXL1* may also function as an oncogene.^41,42^

**Figure 2.**
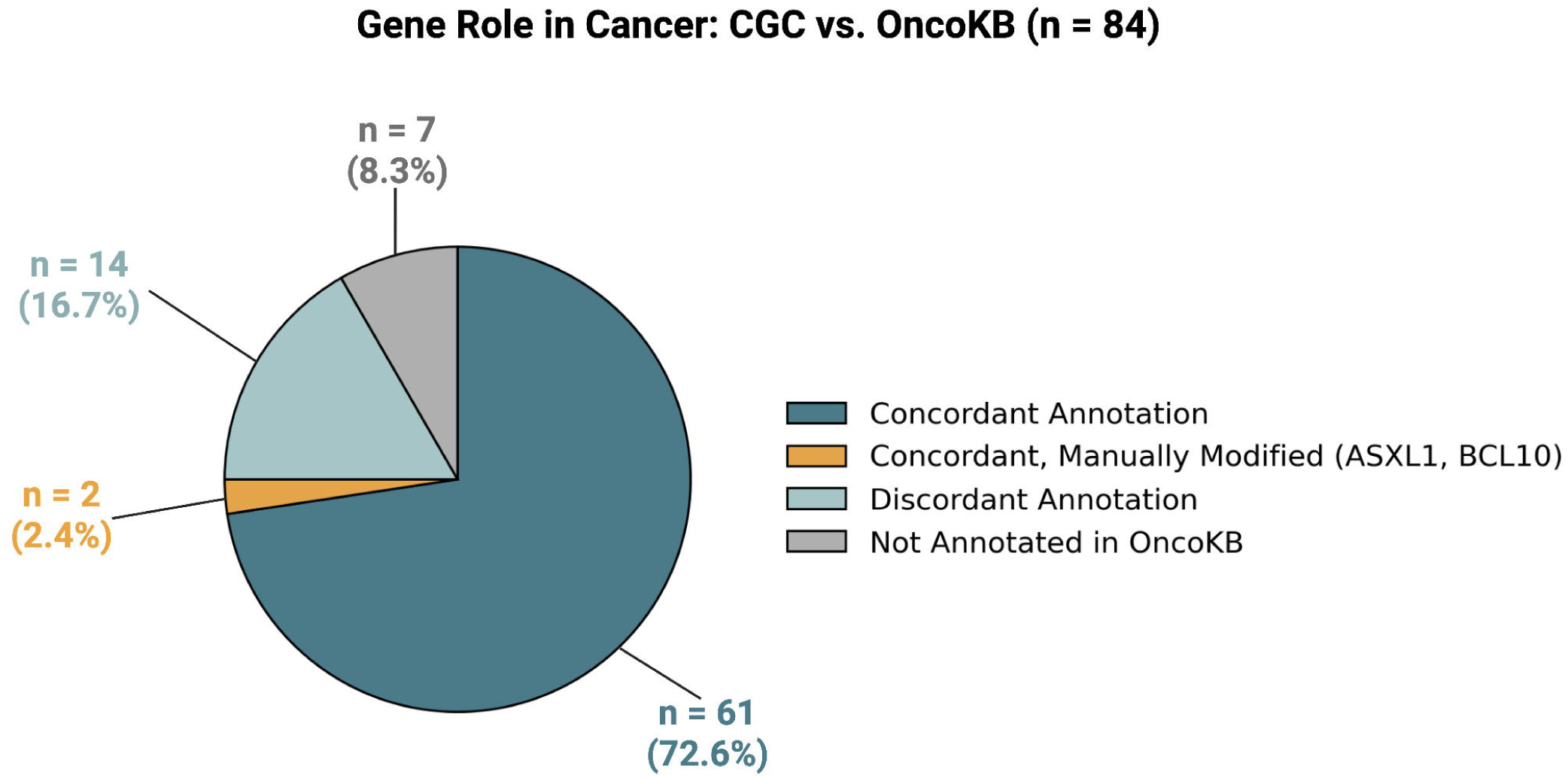
Concordance of gene-role annotations between CGC and OncoKB across 84 genes: 63 concordant, 14 discordant, and 7 without OncoKB annotations. Key manual curation decisions included designating *NPM1* and *RUNX1* as dual-function genes, and reclassifying *ASXL1* and *BCL10* as dual-function despite prior TSG annotations.

After curating the function of the 84 genes, we identified 20 oncogenes and 17 genes with dual roles for downstream analysis, while the remaining 47 genes were TSGs. The oncogenes and dual-function genes harbored 56% (12,416/22,174) of all NMDe variants and were identified in 11,874 unique samples. Of these, ∼59% (7285/12,41) NMDe variants were detected primarily in *CALR* and *NPM1*. The 37 genes did not harbor high-frequency (maximum non-founder subpopulation allele frequency >0.1%) germline PTC-variants in gnomAD (Table S5). The highest allele frequency for an NMDe variant was (*RUNX1)* NM_001754.5:c.1277dup: p.(Arg427AlafsTer174), which is found in 0.1% of the East Asian subpopulation.

### Analysis of Variant Types in Oncogenes and Dual-function Genes

The variant types included nonsense variants (13%, 1,624/12,416), frameshift deletions (∼32%, 3,943/12,416), frameshift insertions (53%, 6,642/12,416) and frameshift indels (2%, 207/12,416) (Figure S2). Notably, *CALR* and *NPM1* show nearl1lexclusive presence of frameshift over nonsense truncating variants (P-value <0.05; supplemental material). In *CALR*, only 4 nonsense variants were seen among 3,467 truncating variants (3,463 were frameshifts). *NPM1* showed a similar pattern (3 nonsense, 3,815 frameshifts).

### Evaluating the C-terminus, Functional Studies and Tumor Phenotype Correlation

To elucidate the mechanisms by which NMDe variants contribute to cancer gene activation, we systematically reviewed 37 genes, examining alterations in the C-terminus, available functional studies, and established disease associations. This analysis identified 11 oncogenes and 4 dual-function genes that exhibited two major patterns of activating NMDe variants (Figure 3; Tables 1 and 2). Below, we summarize published findings that exemplify these mechanisms in oncogenes and dual-function genes, as well as describe NMDe variants causing LoF in a subset of dual-function genes.

**Figure 3.**
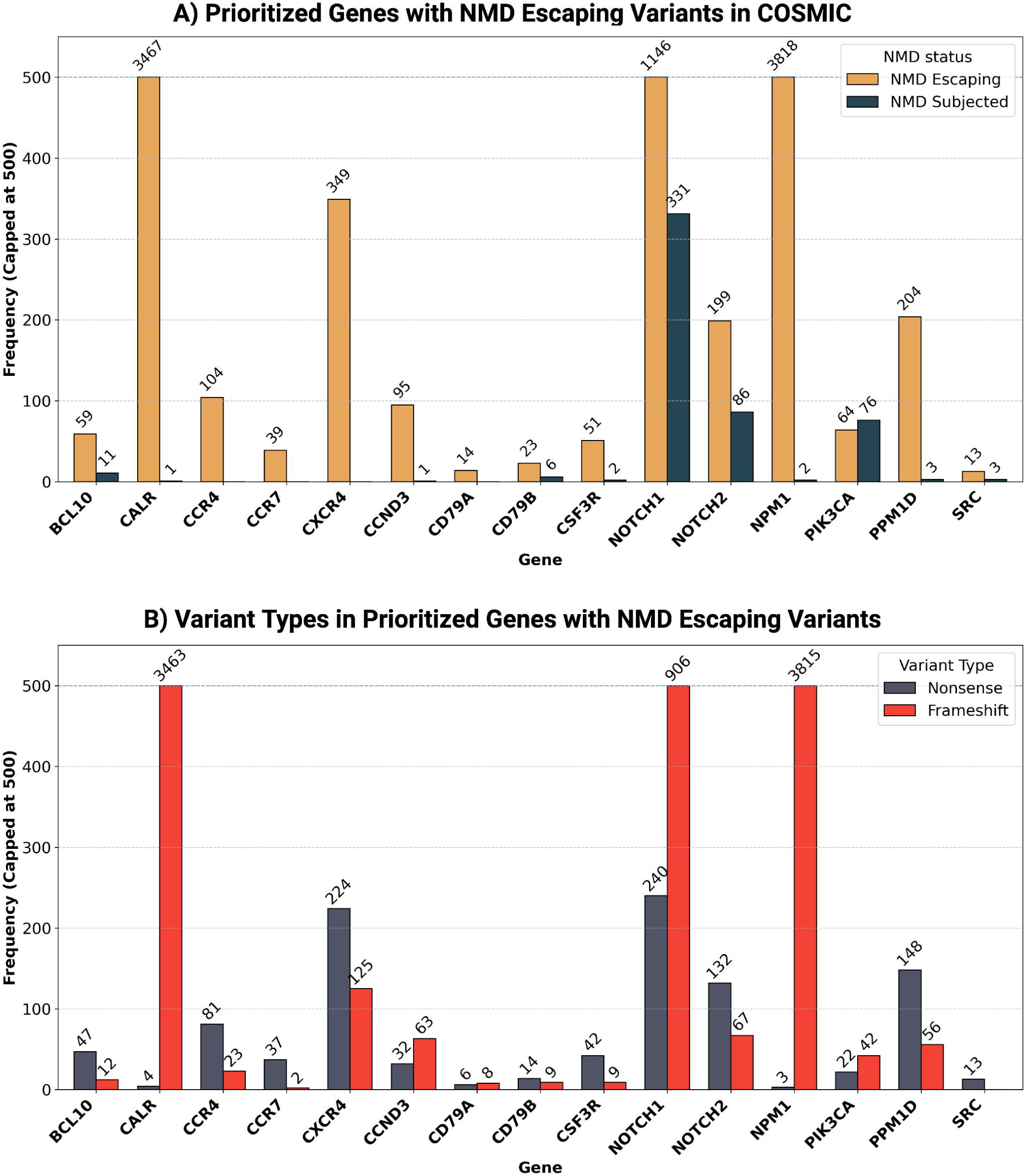
**(A) Prioritized genes (n = 15) with NMDe variants in COSMIC.** Bar chart showing the raw counts of PTC variants classified as NMDe (orange-gold) and NMD-subjected (dark blue) for genes that may harbor activating NMDe variants. *CCR4* and *CXCR4* are predominantly encoded by the second exons and don’t harbor NMD-subjected variants (supplemental material). **(B) Bar chart showing the number of nonsense (dark gray-blue) and frameshift (red) NMDe variants in the same 15 genes curated from the COSMIC database.** Notably, *CALR* and *NPM1* show a marked depletion of nonsense variants. Variant counts are capped at 500 for visualization, with exact counts displayed above each bar.

**Table 1.** Genes demonstrating pattern 1: NMDe variants resulting in an altered C-terminal regulatory region

| Gene | Role in Cancer | NMDe Variant Types | Critical region in C-terminus/<br>Critical Residues | Effects on C-terminus | Signaling Pathway |
| --- | --- | --- | --- | --- | --- |
| CXCR4 | Oncogene | Frameshift/nonsense | Cytosolic region (K308 - S352) encompassing phosphorylation sites (S338/S339) | Attenuated receptor internalization | G protein-coupled receptors |
| CCR4 | Oncogene | Frameshift/nonsense | Phosphorylation sites (T342–T351) | Attenuated receptor internalization | G protein-coupled receptors |
| CCR7 | Oncogene | Frameshift/nonsense | Degradation site (W355) | Attenuated receptor internalization | G protein-coupled receptors |
| *CD79A* | Oncogene | Frameshift/nonsense | ITAM motif (E185–G205) | Attenuated receptor internalization | NF-κB signaling |
| *CD79B* | Oncogene | Frameshift/nonsense | ITAM motif (D193–L213) | Attenuated receptor internalization | NF-κB signaling |
| *CSF3R* | Oncogene | Frameshift/nonsense | Internalization region (S772–F792); degradation region (N818–F836) | Attenuated receptor internalization/ degradation | JAK–STAT pathway |
| *CCND3* | Oncogene | Frameshift/nonsense | PEST degradation sequence<sup>#</sup> (R256–K268); (R271–H291) | Loss of PEST degradation signal; increased stability | CyclinD-CDK4/6-INK4a-Rb pathway |
| *PPM1D* | Oncogene | Frameshift/nonsense | Exon 6 (last exon) | Loss of inhibitory/degradation signal; increased stability and retention of phosphatase domain | DNA-damage response/p53 pathway |
| *SRC* | Oncogene | Frameshift/nonsense | Phosphorylation site (Y530) | Loss of phosphorylation site/reduced downregulation | SRC-Ras-Raf-MEK-ERK1/2 pathway |
| *NOTCH1* | Dual-function | Frameshift/nonsense | PEST degradation sequence<sup>#</sup> (H2507–H2526) | Loss of PEST sequence; prolonged NICD stability and signaling | Notch signaling |
| *NOTCH2* | Dual-function | Frameshift/nonsense | PEST degradation sequence<sup>#</sup> H2412 and H2431 | Loss of PEST sequence; prolonged NICD stability and signaling | Notch signaling |
| *BCL10* | Dual-function* | Frameshift/nonsense | Regulatory region required for inhibition of BCL10 via interaction with MALT1 Ig1–Ig2 (F165–Q208) | Loss of regulatory/inhibitory region; uncontrolled BCL10 polymerization | NF-κB signaling |

**Abbreviations:** Asterisk (\*) indicates gene role in cancer was manually reviewed and modified; ITAM: Immunoreceptor tyrosine-based activation motif. PEST degradation sequence was predicted by EMBOSS epestfind: https://emboss.bioinformatics.nl/cgi-bin/emboss/epestfind; NICD: Notch Intracellular Domain

**Table 2.**
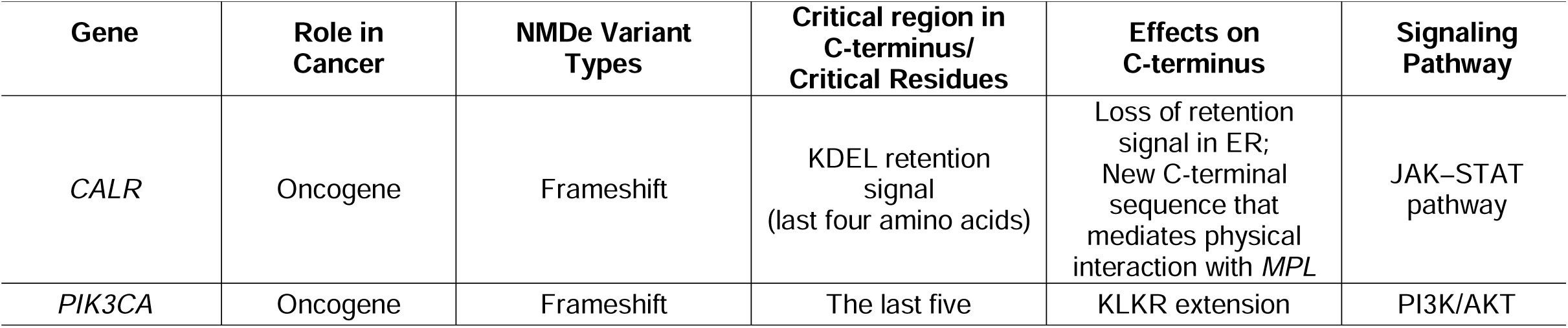
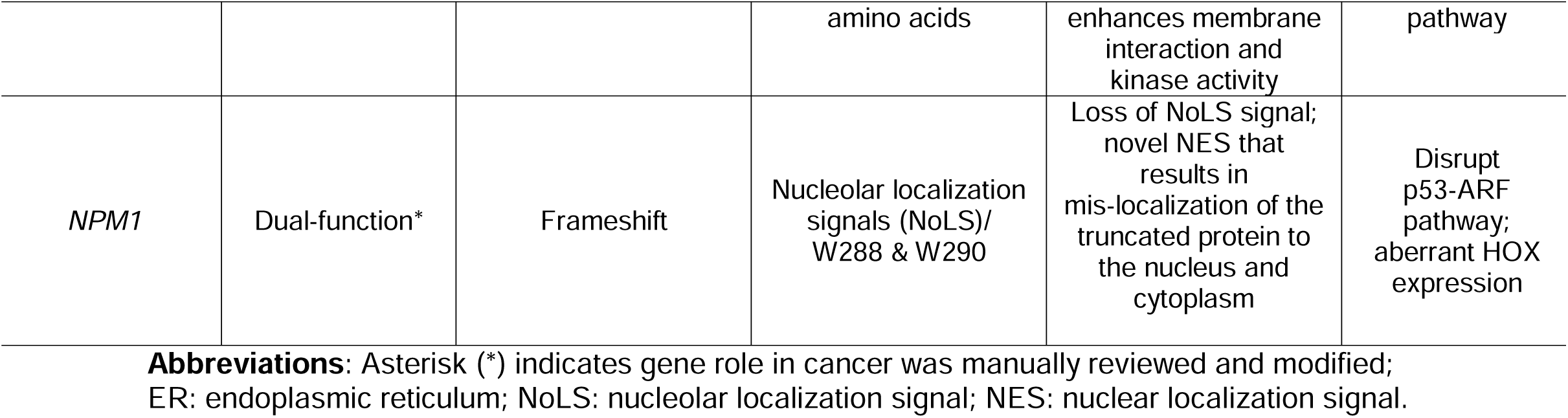
Genes demonstrating pattern 2: NMDe variants create new peptide fragments +/− loss of critical regions resulting in novel/enhanced protein-protein interactions

#### NMDe Variants in Oncogenes

NMDe variants in three chemokine receptors/G protein–coupled receptors (GPCRs) genes were identified, *CXCR4*, *CCR4* and *CCR7,* where NMDe variants resulted in attenuated receptor internalization (Figure S3).^43–48^ In *CXCR4* and *CCR4*, NMDe variants led to truncations that removed key C-terminal phosphorylation sites, including S338/S339 and T342–T351, respectively. Similarly, functional studies revealed that the *CCR7* nonsense variant at W355 in the last exon impairs receptor internalization. NMDe variants in *CD79A* and *CD79B,* which are components of the B-cell receptor (BCR) complex, resulted in loss of immunoreceptor tyrosine-based activation motifs (ITAMs).^49^ Davis and colleagues showed that deletions such as *CD79A* Δ179–226 that extend from the penultimate exon to the last exon and eliminate the ITAMs are associated with increased surface expression of IgM in diffuse large B cell lymphomas (DLBCLs). In *CD79B*, alterations impacting critical tyrosine residues (Y196 or Y207) in the ITAM were also associated with reduced receptor internalization, potentially due to reduced LYN-mediated negative regulation of the BCR complex.^49^ Previous studies showed that truncating *CSF3R* variants that alter the internalization (S772-F792) and/or degradation (N818–F836) signal peptides are predicted to result in attenuated receptor internalization and degradation with a sustained activation of STAT5.^50^ Such alterations in the aforementioned genes are reported in a wide range of hematolymphoid tumors (Supplemental Material).

NMDe variants in *CCND3* are clustered in the last exon (exon 5) that harbor critical residues (T283, P284, I290) that mediate D-type cyclin phosphorylation and stability. These residues lie within a PEST sequence—a stretch of amino acids enriched in proline (<u>P</u>), glutamic acid (<u>E</u>), serine (<u>S</u>) and threonine (<u>T</u>)—that serves as a signal for rapid protein turnover.^14,51^ *CCND3* truncating variants have been reported in mature B cell lymphomas, such as diffuse large B cell lymphomas (DLBCL) and Burkitt lymphoma (BL). *PPM1D* NMDe variants lead to loss of the C-terminal inhibitory/degradation region while retaining phosphatase activity. Consequently, truncated PPM1D functions as a constitutively stable phosphatase that suppresses cellular stress responses by dephosphorylating critical proteins involved in the DNA damage response.^52–55^ *PPM1D* alterations have been described in subsets of CNS and hematolymphoid tumors. Albeit rare, NMDe variants in the SRC proto-oncogene alter the negative regulatory region of the C-terminus that contains a phosphorylation residue (Y530), leading to diminished protein degradation and prolonged activation in a subset of colorectal carcinomas.^56^

*CALR* is a critical diagnostic biomarker in certain myeloproliferative neoplasms.^57^ The *CALR* NMDe variants are predicted to result in the loss of ‘KDEL’ retention signal (last four amino acids) in the C-terminus and introduce a positively charged sequence that mediates physical interaction with MPL (Figure S4).^58^ The *CALR* NMDe variants are predominantly frameshift variants (n= 3467), which generate a motif that shares the terminal amino acid sequence ‘KMSPARPRTSCREACLQGWTEA’ noted in 3455 unique individual samples and previously reported in myeloproliferative neoplasms.^59^ The four nonsense variants detected in *CALR* were in individuals with essential thrombocythemia (Table S2); however, future studies will be critical to elucidate the significance of these nonsense variants.^60–62^

NMDe frameshift variants in *PIK3CA* detected within the last five amino acids led to the generation of a positively charged sequence (‘KLKR’ extension), enhancing membrane interaction and kinase activity.^63–65^ Other truncating *PIK3CA* variants that generate sequences different from the ‘KLKR’ extension require additional functional studies to elucidate their effects. NMDe variants in the remaining nine oncogenes, including *BCL9L*, *CRTC1*, *ERBB2*, *ERG, JUN, MPL, NSD2*, *PIM1* and *SMO*, had limited evidence to determine their effects on the C-terminus.

#### NMDe Variants in Dual-function Genes

The NMDe variants in dual-function genes represent a heterogeneous group in which the resulting effects may depend on the role of the C-terminus or the newly generated peptide sequence. For example, NMDe variants that disrupt C-terminal degradation signals or inhibitory regions can lead to gene activation and enhanced downstream signaling (*NOTCH1*, *NOTCH2*, *BCL10*), whereas those that alter critical regions instead result in loss of function (*BIRC3*, *RUNX1*, *EZH2*) (Supplemental Material). NMDe variants in *NOTCH1/NOTCH2* are associated with loss of the degradation signal (i.e, PEST motif), leading to increased stability of the NOTCH intracellular domain (Figure S5).^66–68^ *NOTCH1* activation is an important prognostic biomarker in chronic lymphocytic leukemia (CLL) and small lymphocytic lymphomas (SLL) and is a key driver in T-lymphoblastic leukemia lymphoma (TLL).^69,70^ Similar *NOTCH2* variants are frequently reported in DLBCL and splenic marginal zone lymphomas (SMZL).^71,72^ *BCL10* NMDe variants lead to the loss of the C-terminal regulatory region, which spans residues 165–208 and is required for inhibition of BCL10 via interaction with MALT1 Ig1–Ig2.^40^ Importantly, these truncating variants still preserve the interaction between the *BCL10* CARD domain and the MALT1 death domain, ensuring robust incorporation of MALT1 into polymerized filaments. Together, these changes lead to potent MALT1 protease activation and sustained NF-κB signaling in a subset of DLBCL.

*NPM1* is a defining biomarker and prognostic indicator in AML.^57^ While precise mechanisms through which truncated *NPM1* contributes to leukemogenesis remain unclear, *NPM1* NMDe frameshift variants are typically characterized by the loss of nucleolar localization signals (NoLS), particularly residues W288 and W290, result in the formation of a novel nuclear export signal (NES) sharing a common terminal peptide sequence ‘VSLRK’, which was noted in 3805 unique samples in COSMIC (Table S2; Figure S5).^73–76^ This results in an aberrant mislocalization of NPM1 to the cytoplasm and destabilization of the p53-ARF tumor suppressor pathway. Additionally, the altered *NPM1* drives leukemogenesis by binding active chromatin and promoting the expression of homeobox (HOX) genes and impairing differentiation.^77^ Rarely, *NPM1* NMDe frameshift variants in the penultimate exon have been described that generate a putative NES with a terminal ‘RRSLIM’ peptide sequence and are associated with cytoplasmic mis-localization of NPM1.^78^ Nonsense (n=3) NMDe variants that result in loss of NoLS alone require further study to elaborate their mechanism and prognostic significance.^79,80^

*ASXL1* NMDe variants are frequently clustered in exon 12 and are associated with the loss of the PHD domain, which has been reported to mediate the interaction with certain modified histone residues.^81,82^ *ASXL1* truncations have also been shown to generate a hyperactive complex with *BAP1*, resulting in upregulation of *HOX* genes through increased deubiquitination of monoubiquitinated histone *H2A* at K119 (H2AK119ub).^81^ Other studies show truncated *ASXL1*, but not wild-type, demonstrated a novel interaction with BET bromodomain-containing protein 4 (BRD4), which is associated with open chromatin and ultimately promotes transcriptional activity genome-wide.^42^ Truncated forms of *ASXL1* typically retain N-terminal residues, including the S201 residue, which is thought to mediate interaction with BRD4. C-terminal NMDe variants in *ASXL1* are noted in a wide range of myeloid neoplasms as well as a subset of CNS tumors.^83,84^

*BIRC3* NMDe variants disrupt the E3 ligase domain, leading to activation of the non-canonical NF-κB pathway in CLL/SLL.^85^ *EZH2* NMDe variants partially disrupt the SET domain, affecting histone methyltransferase activity and causing a DN effect in myeloid neoplasms.^86,87^ *RUNX1* NMDe variants retain the DNA-binding Runt homology domain but lose the ‘VWRPY’ motif that partially alters the transactivation domain, impairing hematopoietic transcriptional regulation and could also be associated with DN effect.^88–92^ *WT1* NMDe variants occurring in exons 9 and 10 were reported in AML and Wilms tumor.^93,94^ The variants are predicted to partially alter the zinc-finger domain, which is essential for DNA/RNA binding.^95–98^ *GATA3* protein C-terminal truncations alter the zinc-finger domain and are most often found in breast tumors and are thought to act as LOF events; however, future studies are warranted to delineate this mechanism.^99,100^ We also detected additional genes where future studies are needed to clarify the effects of NMDe alterations, including *ACVR2A*, *ERBB4*, *MAP3K1*, *NSD1*, *P2RY8*, *TBX3* and *TCF7L2*.

### NMDe Variants in SJCRH Clinical Genomics Cohort

The SJCRH clinical genomics cohort offers a unique opportunity to evaluate NMDe variants in pediatric tumors, enabling assessment of their patterns across tumor types that are underrepresented in COSMIC. Analysis of the 15 prioritized genes (oncogenes and dual-function) identified through reviewing the COSMIC dataset (Table 1 and Table 2) revealed NMDe variants in approximately 3% (113 of 3,492) of unique patient samples, comprising 119 variants across 8 genes (Figure 4; Table S6). The most frequently affected genes were *NOTCH1* (n = 35), *PPM1D* (n = 24), *CCND3* (n = 22), *NPM1* (n = 19), *CXCR4* (n = 9), and *CSF3R* (n = 8), with single occurrences in *CD79B* and *NOTCH2*. No NMDe variants were detected in the remaining seven genes identified from the COSMIC analysis.

**Figure 4.**
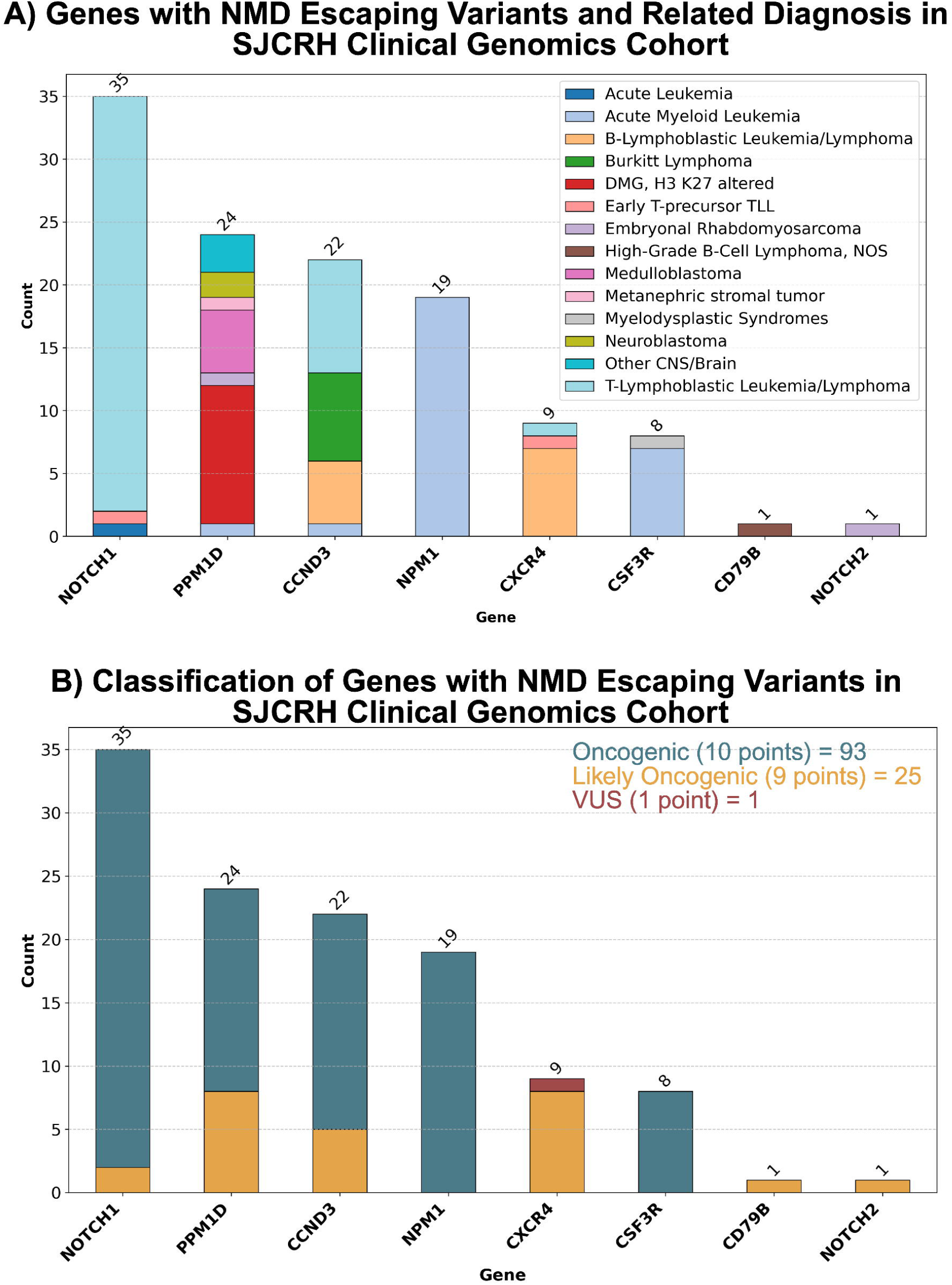
Interrogation of prioritized genes in the SJCRH clinical genomics cohort. (A) The highest frequencies were observed in *NOTCH1*, *PPM1D*, *CCND3*, *NPM1*, *CXCR4*, and *CSF3R*. Bars are colored by tumor type. (B) Classification of NMDe variants in oncogenes and dual-function genes based on the application of OVS1_Very Strong, OP3_Supporting and OP4_Supporting evidence codes. Variants were classified as oncogenic (n = 93), likely oncogenic (n = 25), or variant of uncertain significance (n = 1)

Leveraging the SJCRH cohort, we attempted to combine the classification criteria (OVS1_Very Strong, OP3_Supporting and OP4_Supporting, see Methods) from the NMD analysis with available clinical knowledge. OVS1_Very Strong (8 points) was applied to NMDe variants in 7 genes that disrupted critical regulatory (inhibitory/degradation) regions, as well as to *NPM1* (Table 3). All *NPM1* NMDe variants were frameshift alterations (n= 19), which are predicted to result in loss of the W288 and W290 residues and generate a novel C-terminal peptide fragment containing a nuclear export signal (NES) that terminates with ‘VSLRK’ (n = 18) and one variant predicted to create a NES ending with ‘RRLSIM’. OP4_Supporting (1 point) was applied due to the absence of high-frequency germline PTC variants (>0.1%) in gnomAD.

**Table 3.** Summary of Variant Classifications.

| Gene<br>(number of<br>variants) | OVS1 Evidence<br>(8 points) | OP4<br>Evidence *<br>(1 point) | OP3 Evidence<br>(1 point)<br>(Tumor type-specific<br>study) | Total<br>Points<br>(Frequency) | Final<br>Classificatio |
| --- | --- | --- | --- | --- | --- |
| *CCND3*<br>(n=22) | Loss of PEST degradation<br>sequence (R271-H291) | Absent or<br>< 0.1% in<br>gnomAD | TLL (n=9); BL (n=7);<br>AML with *KMT2A*<br>rearrangement (n=1) | 10 points (n=17);<br>9 points (n=5) | Oncogenic;<br>Likely<br>Oncogenic |
| *CD79B*<br>(n=1) | Loss of ITAM motif<br>(D193–L213) | < 0.1% in<br>gnomAD | NA | 9 points (n=1) | Likely<br>Oncogenic |
| *CSF3R*<br>(n=8) | Loss of receptor Internalization<br>region<br>(S772-F792);<br>degradation region<br>(N818–F836) | Absent or<br>< 0.1% in<br>gnomAD | AML (n=7);<br>MDS (n=1) | 10 points (n=8) | Oncogenic |
| *CXCR4*<br>(n=9) | Altering cytosolic region (K308 -<br>S352) mediating for receptor<br>internalization | Absent or<br>< 0.1% in<br>gnomAD | NA | 9 points (n=8);<br>1 point (n=1) | Likely<br>Oncogenic;<br>VUS |
| *NOTCH1*<br>(n=35) | Loss of the PEST degradation<br>sequence (H2507–H2526) | Absent or<br>< 0.1% in<br>gnomAD | TLL (n=33) | 10 points (n=33);<br>9 points (n=2) | Oncogenic;<br>Likely<br>Oncogenic |
| *NOTCH2*<br>(n=1) | Loss of the PEST degradation<br>sequence (H2412 and H2431) | Absent or<br>< 0.1% in<br>gnomAD | NA | 9 points (n=1) | Likely<br>Oncogenic |
| *NPM1*<br>(n=19) | Loss of nucleolar localization<br>signals (NoLS) (W288 and/or<br>W290); newly generated sequence<br>NES (CLAVEEVSLRK) or<br>(RRLSIM) | Absent or<br>< 0.1% in<br>gnomAD | AML<br>(n=19) | 10 points (n=19) | Oncogenic |
| *PPM1D*<br>(n=24) | NMDe in the last exon<br>(exon 6) that mediates<br>proteasomal degradation | Absent or<br>< 0.1% in<br>gnomAD | DMG, H3 K27-altered (n=11);<br>MB (n=5) | 10 points (n=16);<br>9 (n=8) | Oncogenic;<br>Likely<br>Oncogenic |

**Abbreviations:** AML: Acute Myeloid Leukemia; BL: Burkitt Lymphoma; DMG: Diffuse Midline Glioma; TLL: T-Lymphoblastic Leukemia/Lymphoma; ETP ALL: Early T-precursor lymphoblastic Leukemia/Lymphoma; MDS: Myelodysplastic Syndromes; NES: nuclear export signal. Asterisk denotes that OP4\_supporting (1 point) evidence was applied to variants either absent or less than 0.1% in gnomAD (v4.1.0).

The (*CXCR4*) NM_003467.3 c.240_241insGAGTGGGGGGG p.(Ser81GlufsTer91) variant, identified in a patient with B-lymphoblastic leukemia/lymphoma, was regarded as of uncertain significance (VUS). This variant is predicted to truncate a major portion of the CXCR4 seven-transmembrane domain, resulting in a protein length change of 78% (Figure 5). Due to the magnitude of truncation and uncertain functional consequences, OVS1 was not applied and the clinical significance of this variant remains to be determined.

**Figure 5.**
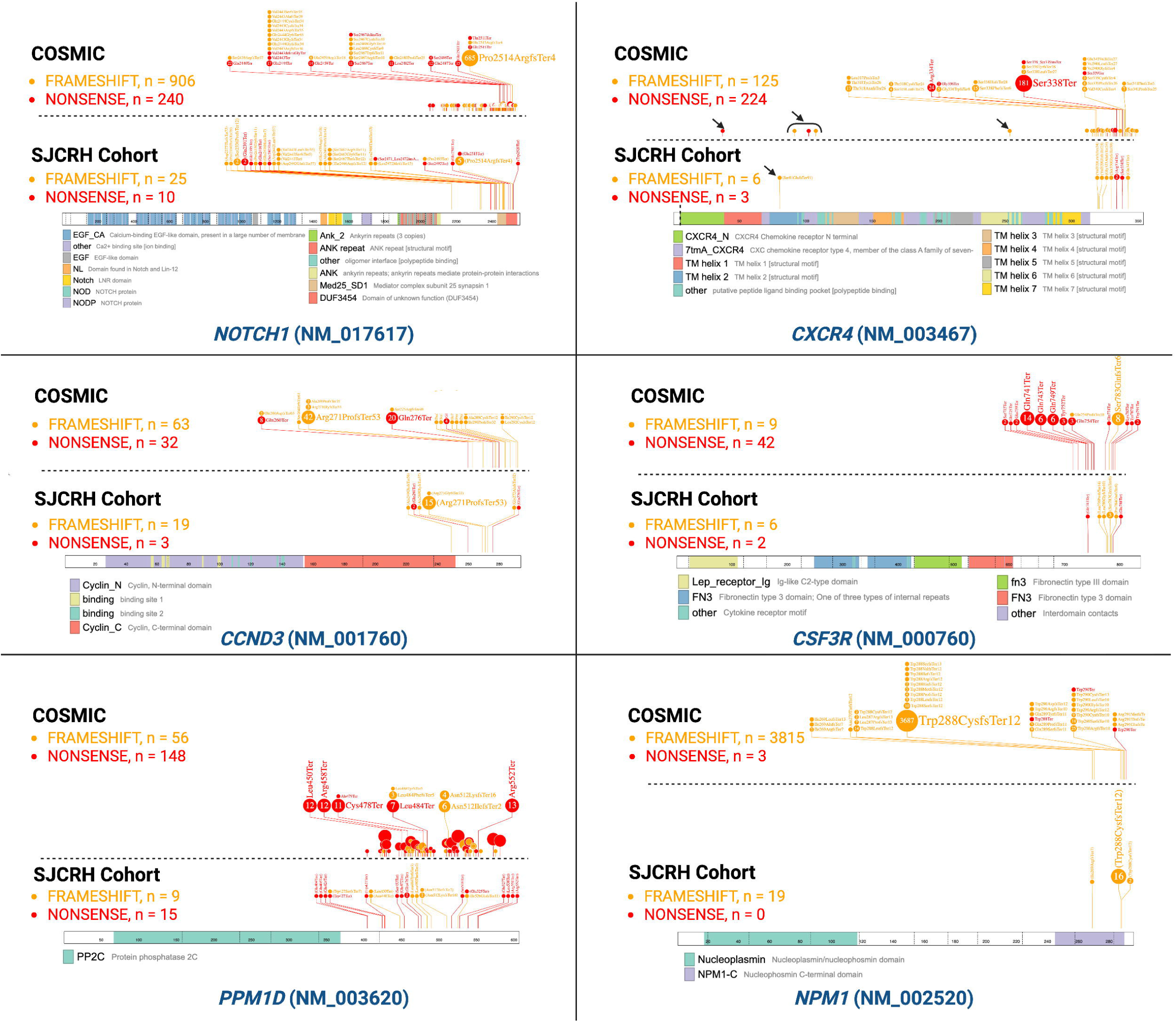
Distribution of NMDe variants in the SJCRH Clinical Genomics Cohort compared to COSMIC. ProteinPaint plots show the positional distribution of NMDe variants across six representative genes (*NOTCH1, CXCR4, CCND3, CSF3R, PPM1D*, and *NPM1*). For each gene, COSMIC data (top panel) are displayed alongside variants identified in the St. Jude Clinical Genomics Cohort (bottom panel). Frameshift and nonsense variants are indicated in orange and red, respectively. The vertical dashed line in *CXCR4* demarcates exons 1 and 2; arrows highlight five NMDe variants in COSMIC and one in the SJCRH cohort predicted to truncate the seven-transmembrane domains, for which the functional significance remains uncertain.

In total, 118 NMDe variants found in the SJCRH cohort met criteria as activating NMDe variants (as defined in Table 1; Table 2) and meeting the criteria for being likely oncogenic (9 points). Furthermore, 93 variants across five genes (*CCND3*, *CSF3R*, *NOTCH1*, *NPM1* and *PPM1D*) were detected in a relevant tumor type (Table 3), and application of OP3_Supporting (1 point) contributed to oncogenic classification (10 points) for these variants. In combination, the application of OVS1_Very Strong, OP3_Supporting, and OP4_Supporting for NMDe variants in oncogenes and dual-function genes resulted in the classification of 93 variants as oncogenic, 25 as likely oncogenic, and one remained a VUS (Figure 4).

## Discussion

We conducted a comprehensive screening of the COSMIC database, one of the largest and most detailed repositories of cancer data, to flag and characterize genes that may harbor somatic variants predicted to escape NMD and promote gene activation. While we analyzed raw variant counts from COSMIC, approximately 36% of PTC-variants were predicted to escape NMD, which aligns with prior reports of this mechanism.^101,102^ We further focused on well-established genes harboring at least ten NMDe variants and for which gene-level and variant-level functional studies were available. This approach enabled us to prioritize 15 genes (oncogenes or dual-function genes) that harbored two distinct patterns of NMDe variants that can result in gene activation.

The first pattern (Table 1) comprises NMDe variants that disrupt C-terminal regulatory regions that play roles in controlling protein inhibition and/or degradation. By removing critical residues (e.g., phosphorylation sites or PEST motifs), these alterations can result in enhancing downstream signaling by impaired receptor internalization/degradation in several cell membrane receptors or increased intracellular protein stability. Notably, in *BCL10*, loss of the C-terminal inhibitory region results in uncontrolled BCL10 polymerization.^40^ The second pattern (Table 2) is exclusively driven by frameshift NMDe variants that generate novel peptide sequences. The resulting peptide sequences can mediate protein interactions that enhance existing ones (*PIK3CA*) or produce neomorphic effects (*CALR*, *NPM1*), in addition to causing loss of critical protein regions (e.g., ‘KDEL’ retention signal in *CALR* and NoLS in *NPM1*).^58,77^ For this pattern, it is important to carefully consider the spectrum of reported variant types, especially comparing frameshift variants with nonsense variants, the latter would be expected to result in protein truncation without introducing novel peptide fragments. For frameshift variants, examination of the sequence of peptide fragments (e.g., ‘VSLRK’ in *NPM1*) generated by the NMDe variants could help verify oncogenic effects.

Subsequent interrogation of the SJCRH clinical genomics dataset revealed that ∼3% (113/3,492) of unique patient samples from pediatric patients carried 119 NMDe variants. While we acknowledge subjectivity in applying evidence codes and strength, activating NMDe variants were identified in ∼99% (118/119) of the analyzed variants and met the criteria for at least a likely oncogenic classification. Collectively, analysis of COSMIC and SJCRH cohorts underscores the need for a comprehensive framework that considers the biological roles of genes in cancer more broadly (TSGs vs. oncogenes vs. dual-function genes), variant types, evaluates the specific functions of C-terminal regions removed and assesses the effects of newly generated protein sequences. This comprehensive framework can also shed light on our understanding of the variable consequences associated with somatic NMDe variants.

Our analysis highlights additional key challenges and important considerations in the interpretation of somatic NMDe variants. We observed discordance in gene function annotations between OncoKB and CGC in ∼17% (14/84) of COSMIC-screened genes, highlighting the need for a standardized framework of terminology and criteria to define a gene’s role in cancer. The positional context of activating NMDe variants was also critical, particularly in single coding exon genes (*CCR4*) or genes largely encoded by the second exon (*CXCR4*), where N-terminal variants truncate substantial protein regions. NMDe variants resulting in LoF via dominant-negative effects should also be evaluated, rather than restricting assessment to variants that alter critical regions, as illustrated by *EZH2*.

There are several important limitations to this study. Because the COSMIC targeted screening dataset aggregates data from heterogeneous studies, raw variant counts are influenced by factors such as panel design, per-gene testing frequency, tumor type distribution, and study-specific reporting biases. We acknowledge that other genes (e.g., *CCND2*) with fewer than ten NMDe somatic variants in COSMIC harbor biologically and clinically relevant activating NMDe variants.^103^ We didn’t evaluate other alternative ‘non-canonical’ rules that mediate the evasion of NMD.^5^ We did not assess the effects of splicing variants in the last intron on evading the NMD. Caution is warranted before generalizing our findings, for example in the case of *ASXL1,* which demonstrates considerable biological complexity. We exclusively addressed the somatic interpretation of NMDe variants and did not consider germline NMDe variants although they likely also pay a role in hereditary disorders. Future large-scale efforts might be warranted to assess the ClinGen framework for the evaluation of germline NMDe variants in genes not characterized by a LoF disease mechanism. For instance, germline NMDe variants could result in gene activation by removing the C-terminal inhibitory/degradation region in a subset of autosomal dominant genes, such as *CCND2* in megalencephaly-polymicrogyria-polydactyly-hydrocephalus syndrome 3 (MIM: 615938), *CXCR4* in warts, hypogammaglobulinemia, infections and myelokathexis (MIM: 193670), *EPOR* in familial erythrocytosis (MIM: 133100) and *NOTCH2* in Hajdu-Cheney syndrome (MIM: 102500).^4,104–108^

In conclusion, NMDe is a critical disease mechanism that activates multiple actionable oncogenes and dual-function genes by removing a regulatory/inhibitory C-terminal region or through frameshift variants introducing novel peptide sequences. Evaluating the gene function, altered C-terminal regions, newly generated protein sequences and variant types is indispensable for the comprehensive interpretation of somatic NMDe variants. This comprehensive approach can elucidate the diverse functional consequences associated with NMDe variants and underscores the need to expand assessment of NMDe in the joint recommendations beyond TSGs. Future studies are necessary to further refine the evidence strength for interpreting NMDe variants in oncogenes and dual-function genes.

## Data Availability

All relevant data produced in the present work are contained in the manuscript and supplemental files

## Web resources

COSMIC, https://cancer.sanger.ac.uk/cosmic

EMBOSS epestfind, https://emboss.bioinformatics.nl/cgi-bin/emboss/epestfind

gnomAD, https://gnomad.broadinstitute.org/

HGVS, https://hgvs-nomenclature.org/

National Comprehensive Cancer Network, https://www.nccn.org/

OncoKB, https://www.oncokb.org/

ProteinPaint, https://proteinpaint.stjude.org/

VariantValidator, https://www.variantvalidator.org/

VEP, https://useast.ensembl.org/info/docs/tools/vep/index.html

## Supplemental information

**Table S1.** Variants Excluded from COSMIC

**Table S2.** Variants Included in COSMIC Analysis

**Table S3.** Manual Gene Function Curation

**Table S4.** Aggregate Summary of Variants

**Table S5.** gnomAD Variants in Curated Genes

**Table S6.** SJCRH Verification Cohort

## Data and code availability

All data supporting the findings of this study are available within the article and its supplemental information. COSMIC dataset analyzed was obtained from public repositories. A custom script to determine the NMD status is available at https://github.com/meldomer.

## Declaration of interests

S.E.P. reports financial support from the National Human Genome Research Institute and serves on scientific advisory panel of Baylor Genetics Laboratories, LLC. All other authors declare no competing financial interests.

## Acknowledgments

This work was funded by the American Lebanese and Syrian Associated Charities of SJCRH. S.E.P. received funding from 2 U24HG009649. Preliminary findings of this work were presented at the Association of Molecular Pathology (AMP) 2025 Annual Meeting & Expo.

## Author contributions

Conceptualization, M.K.E.; supervision: M.K.E., S.E.P., formal analysis, M.K.E.; statistical analysis: J.L., L.T.; data curation, M.K.E., P.R.B., K.D.J, M.N., J.N.; visualization, M.K.E.; writing – original draft, M.K.E.; writing – review & editing, K.D.J, M.N., J.N., R.D.K, M.R.W., J.L., L.W., J.M.K., L.T., S.E.P., P.R.B.

## Notes

### Author Declarations

The study was reviewed by the Institutional Review Board deemed exempt and approved (IRB 25-2016); informed consent was not required per the St Jude Children Research Hospital IRB. All data were deidentified

