## Supplemental Material for "Analysis and Interpretation of Somatic NMD-Escaping Variants in Oncogenes and Dual-Function Genes across Adult and Pediatric Cancer Cohorts"

**Supplemental Methods**

**Analysis of Variant Types in COSMIC**

For each gene, we evaluated the other 36 genes using a negative binomial regression model adjusted for the number of NMD-escaping (NMDe) variants and CDS length. This model was then used to estimate the expected nonsense variant count for the gene of interest. Finally, the observed nonsense count was compared with the expected distribution under the negative binomial model to derive a p-value for depletion.

**Supplemental Results**

**NMDe variants in Oncogenes**

The MANE Select transcript (NM_003467) of *CXCR4* (C-X-C motif chemokine receptor 4) consists of two exons. The first exon encodes five amino acids, while the second exon accounts for the remaining 99% of the protein, encoding 349 amino acids. *CXCR4* NMDe variants are typically clustered in the C-terminal regulatory cytosolic region (K308 - S352), resulting in the loss of critical phosphorylation residues necessary for interaction with β-arrestins, which mediate receptor internalization.^1-5^ In the analyzed COSMIC dataset, NMDe variants (n = 310) were clustered between T311 and S339 and are predicted to disrupt key residues, such as S338/S339, which are essential for receptor internalization.^6,7^ NMDe variants occurring downstream of S338/S339 were also observed, affecting other serine residues (346, 347, 348, 351, and 352) that may also impact receptor regulation.^7-11^ *CXCR4* NMDe variants were identified in lymphoid neoplasms, particularly lymphoplasmacytic lymphoma/Waldenström macroglobulinemia (n=300). Notably, *CXCR4* NMDe variants have therapeutic relevance, influencing responses to certain Bruton tyrosine kinase (BTK) inhibitors in lymphoplasmacytic lymphoma.^5^ Five variants were noted between residues 38 and 195 and were found in diffuse large B cell lymphomas (DLBCLs).^12^ While these N-terminal NMDe variants are predicted to alter critical residues, the variants are predicted to truncate a major portion of the *CXCR4* seven-transmembrane domains and future studies are needed to elucidate the significance of those alterations.

NMDe variants in *CCR4* (C-C motif chemokine receptor 4) and *CCR7* (C-C motif chemokine receptor 7) have been reported in adult T-cell lymphoma/leukemia.^13,14^ NMDe variants in *CCR4* and *CCR7* have been shown to attenuate receptor internalization in response to ligand stimulation, resulting in persistently elevated surface expression of the receptor, even in the absence of ligand, as well as augmented ligand-induced chemotaxis.^13^ *CCR4* (NM_005508) consists of two exons, with the first exon being non-coding. Approximately 99% (102/104) of *CCR4* NMDe variants are located in the C-terminal region between residues 323 and 347, a region that harbors critical phosphorylation sites (between residues T342–T351) necessary for mediating receptor internalization.^13,14^ *CCR7* (NM_001838) consists of three exons, with the first exon also being non-coding. *CCR7* NMDe variants (n= 39) cluster in the third exon between residues 341 and 355. Functional studies demonstrated that the *CCR4* p.(Y331*) and *CCR7* p.(W355*) nonsense variants impair receptor internalization.^13^

*CSF3R* (colony stimulating factor 3 receptor), a member of the class I cytokine receptor (hematopoietin receptor) superfamily, contained 51 NMDe variants.^15,16^ Importantly, a subset of *CSF3R* NMDe variants located between E700-T738 reduced receptor internalization but, on their own, did not activate STAT5.^16^ These upstream truncations gained transforming capacity when combined with the membrane-proximal activating variant T618I, underscoring the requirement for intact STAT5 activation in oncogenic signaling. In COSMIC, 48 of 51 NMDe variants were located downstream of T738. *CSF3R* NMDe variants were reported in acute myeloid leukemia (AML), chronic neutrophilic leukemia, and atypical chronic myeloid leukemia.^17,18^

*CCND3* (cyclin D3) belongs to the cyclin D family, which activates CDK4 (cyclin-dependent kinase 4) or CDK6 (cyclin-dependent kinase 6).^19^ The PEST sequence is predicted to span residues R256–K268 and R271–H291 in exon 5 (predicted by EMBOSS epestfind: <https://emboss.bioinformatics.nl/cgi-bin/emboss/epestfind>). *CCND3* truncating variants (n = 95) are expected to result in loss of critical phosphorylation sites within the PEST degradation sequence, leading to increased protein stability.^19,20^ In addition to Burkitt Lymphoma and DLBCLs, *CCND3* NMDe variants have also been reported in a subset of AMLs.^21,22^ *PPM1D* (protein phosphatase, Mg²⁺/Mn²⁺–dependent 1D), also known as Wip1 (wild-type p53-induced phosphatase 1), harbors NMDe variants (n = 204) clustered in exon 6.^23^ *PPM1D* variants are reported in a variety of neoplasms, including diffuse midline glioma and clonal hematopoiesis after exposure to chemotherapy.^23-26^

CALR (calreticulin) primarily functions in protein folding and calcium homeostasis. *CALR* NMDe variants also result in the removal of the ‘KDEL’ endoplasmic reticulum retention signal and the newly formed basic peptide tail acquires specific affinity for the thrombopoietin receptor (MPL), promoting stable interactions that facilitate MPL trafficking to the cell surface and constitutive activation of the JAK-STAT signaling pathway.^27-30^ The *CALR* mutant C-terminus enhances the binding of the CALR N-terminus to immature N-glycans by increasing the accessibility of its N-terminal domain. *CALR* NMDe variants are detected in myeloproliferative neoplasms, particularly in essential thrombocythemia (n=2217) and primary myelofibrosis (n=1145) (Table S2; S6).^31^

**NMDe variants in dual-function genes**

NMDe variants in *NOTCH1* (notch receptor 1) and *NOTCH2* (notch receptor 2) are predicted to disrupt the PEST sequence. Functional studies have shown that loss of this region increases the half-life of the Notch intracellular domain by impairing PEST-mediated degradation.^32,33^ In *NOTCH1*, 1,146 NMDe variants have been identified, of which 1,142 are predicted to affect the PEST sequence spanning residues H2507–H2526. These variants are frequently reported in chronic lymphocytic leukemia/small lymphocytic lymphoma (CLL/SLL), T-acute lymphoblastic leukemia/lymphoma (T-ALL), and adenoid cystic carcinoma.^34-37^ Of the detected NMDe variants in *NOTCH2* (n = 199), 195 are predicted to alter the *NOTCH2* PEST sequence spanning residues H2412-H2431. *NOTCH2* NMDe variants are recurrent in mature B cell lymphomas, including DLBCLs and splenic marginal zone lymphomas.^12,38-40^ Conversely, somatic NMD-subjected/inactivating variants in *NOTCH1* (n = 139) and *NOTCH2* (n = 36) are well documented in certain squamous cell carcinomas.^41,42^

NMDe variants in *BCL10* (BCL10 immune signaling adaptor) frequently truncate the C-terminus, removing residues 165–208 that normally mediate an inhibitory interaction with the *MALT1* immunoglobulin-like (Ig1–Ig2) domain.^43^ Loss of this regulatory region abrogates the negative feedback that restrains BCL10 polymerization, leading to constitutive filament assembly. As a result, these *BCL10* truncating variants result in potent MALT1 protease activation and downstream NF-κB signaling. Truncations downstream to Q208 truncate the extreme C-terminal Ser/Thr-rich tail; however, the functional significance of this region is yet to be determined. *BCL10* NMDe alterations are primarily found in DLBCLs (n=53). *BCL10* NMD-triggering variants in COSMIC were reported in epithelial tumors, B-lymphoblastic leukemia/lymphoma, breast implant-associated anaplastic large cell lymphoma *in situ*, and angioimmunoblastic T-cell lymphoma.

*BIRC3* (baculoviral IAP repeat containing 3) NMDe variants are associated with the loss of E3 ubiquitin ligase activity of the RING domain that negatively regulates MAP3K14, which is required for activation of the non-canonical NF-κB pathway.^44,45^ Both *BIRC3* NMDe (n = 68) or NMD-triggering (n= 36) variants occur in CLL/SLL. Conversely, overexpression of *BIRC3* has been reported in a subset of gliomas.^46^ *RUNX1* (RUNX family transcription factor 1), a pivotal transcription factor essential for hematopoietic differentiation, functions as both a transcriptional activator and repressor.^47,48^ *RUNX1* NMDe variants lead to altered transactivation and/or affect the conserved VWRPY motif, likely resulting in a dominant negative effect.^49-53^ *RUNX1* NMD escaping variants were noted in individuals with myeloid neoplasms (n=173), including AMLs and myelodysplastic syndrome. Nevertheless, overexpression of *BIRC3* and *RUNX1* might have a distinct disease mechanism compared to protein depletion or loss of critical regions.^46,54^

NMDe variants in *EZH2* (enhancer of zeste 2 polycomb repressive complex 2 subunit), a catalytic subunit of the polycomb repressive complex 2 (PRC2), are predicted to partially disrupt the SET domain that exhibits histone methyltransferase activity.^55,56^ Notably, functional studies have shown that C-terminal NMDe variants in the *EZH2*, including those downstream to the SET domain, such as NM_004456.5:c.2231_2232insGGCTGACCGGCA p.(Ile744MetfsTer23), are associated with a dominant negative effect and reduction of the H3K27 trimethylation.^55^ EZH2 NMDe alterations were noted in several myeloid neoplasms, including AMLs (n=12).^57^ Moreover, the case of *EZH2* exemplifies how an epigenetic regulator can act as an oncogene in some contexts and a tumor suppressor in others. In mature B-cell lymphomas, activating amino acid substitutions in *EZH2* (at Y641 and other residues) increase its methyltransferase activity, leading to excessive silencing of differentiation genes and resulting in a block in B-cell maturation.^58,59^ EZH2 inhibitors are being used clinically to treat such lymphomas. In contrast, in T-ALL and some myeloid malignancies, *EZH2* or other PRC2 components are often deleted or inactivated.^55,56^

**Supplemental Figures**

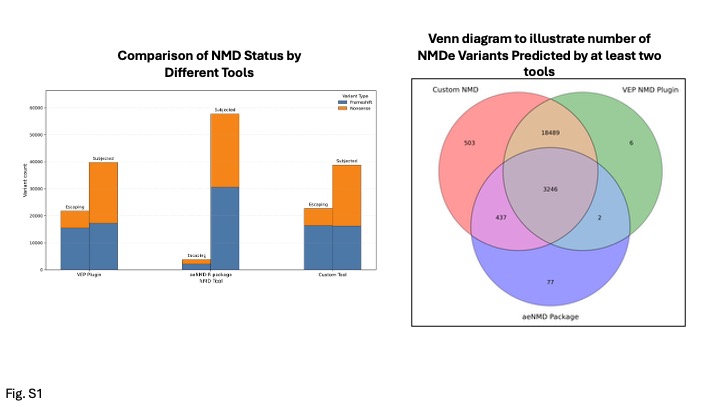

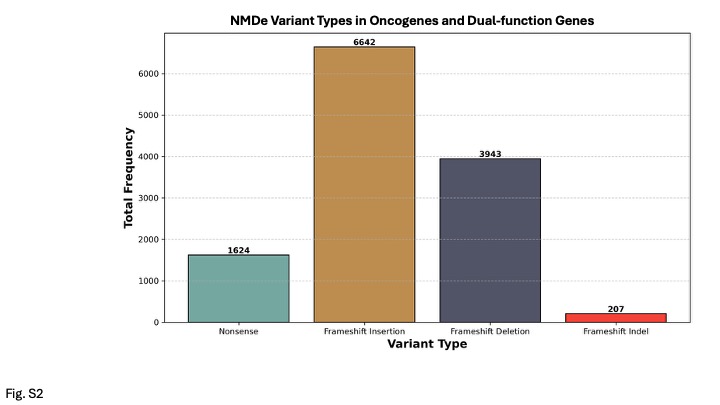

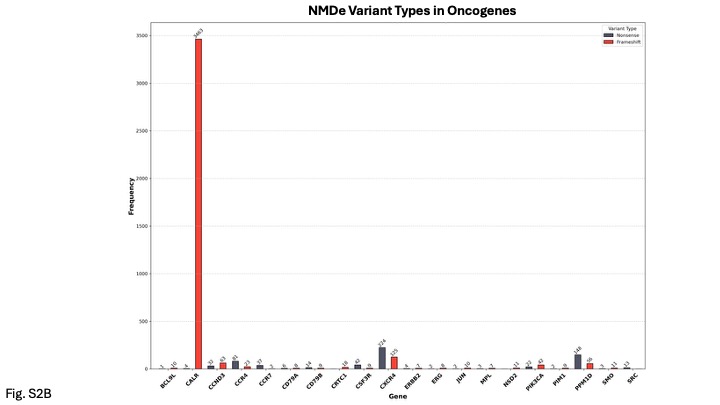

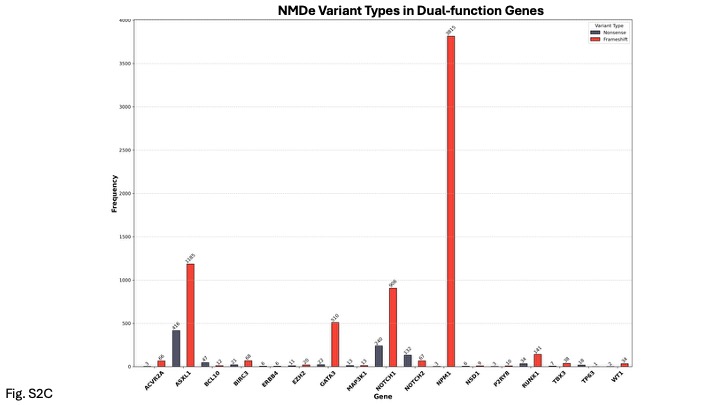

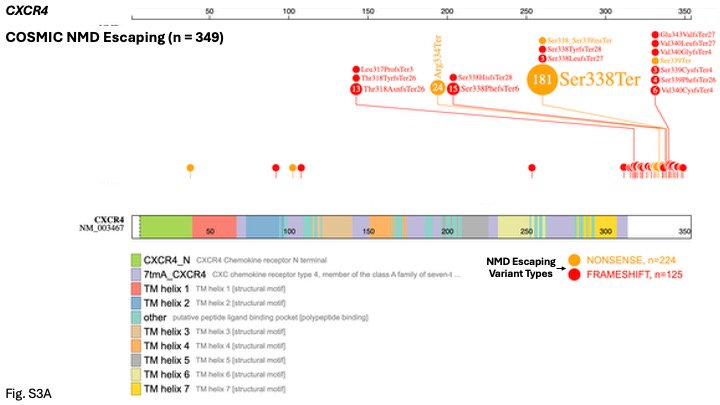

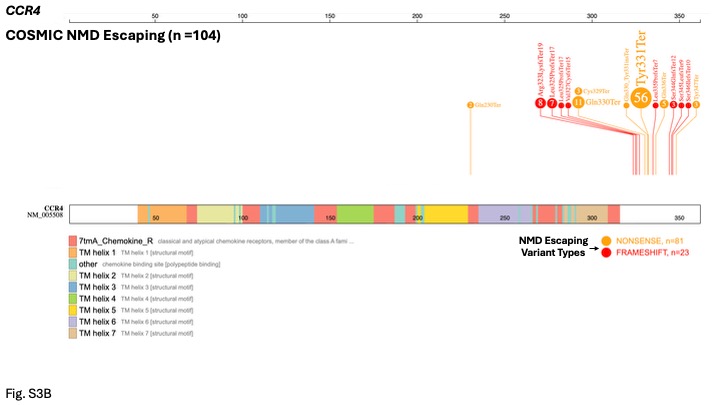

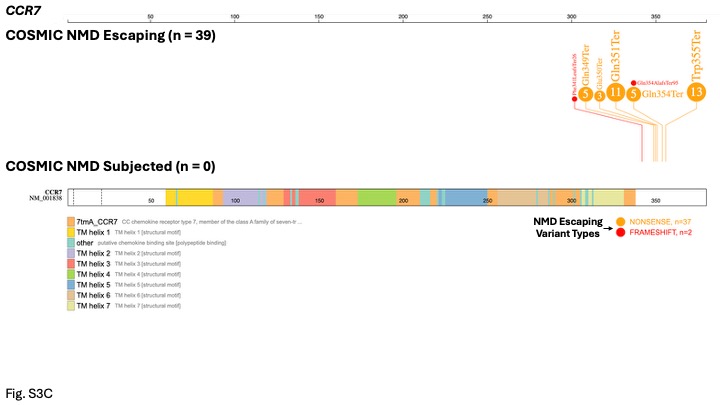

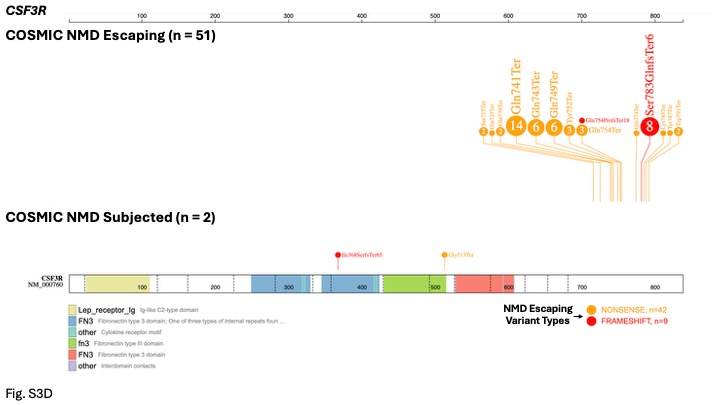

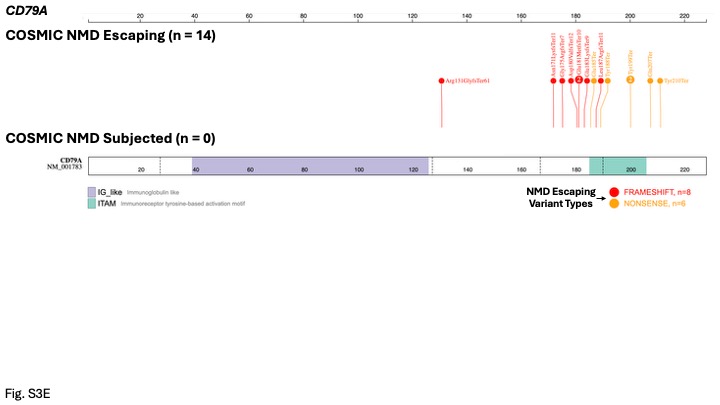

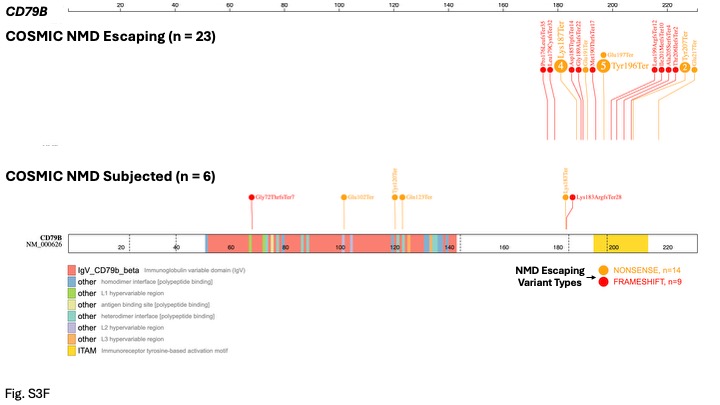

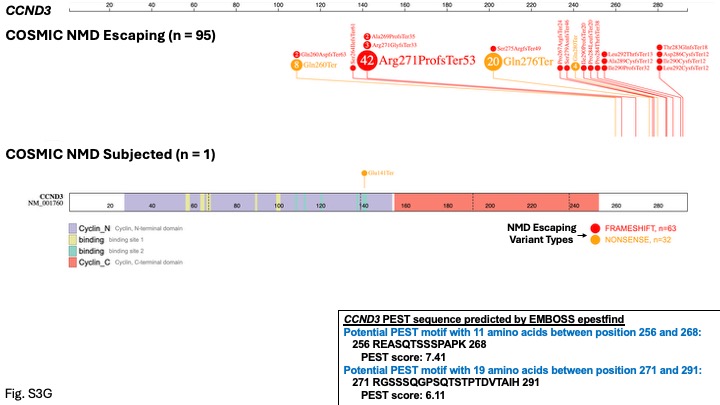

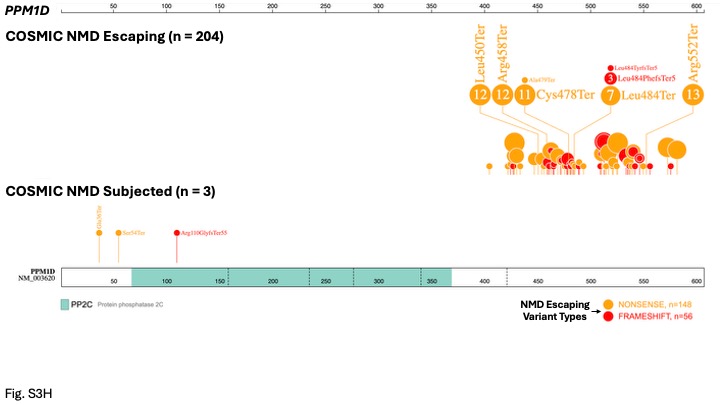

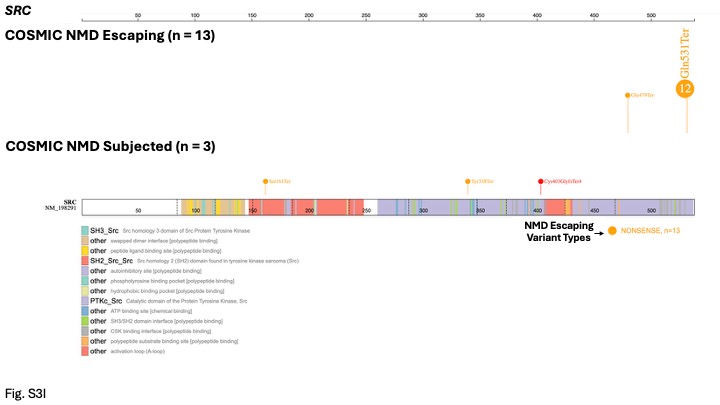

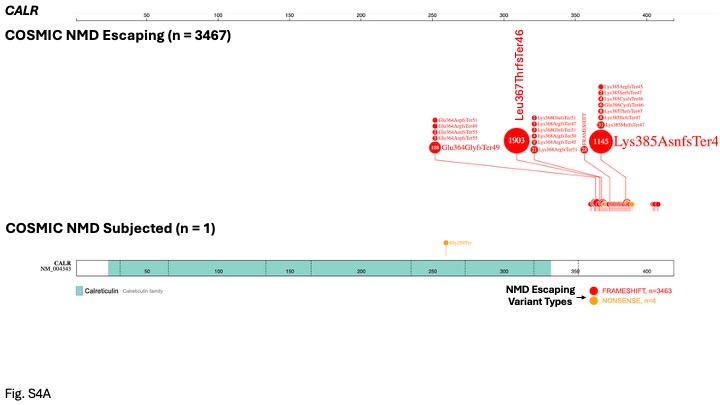

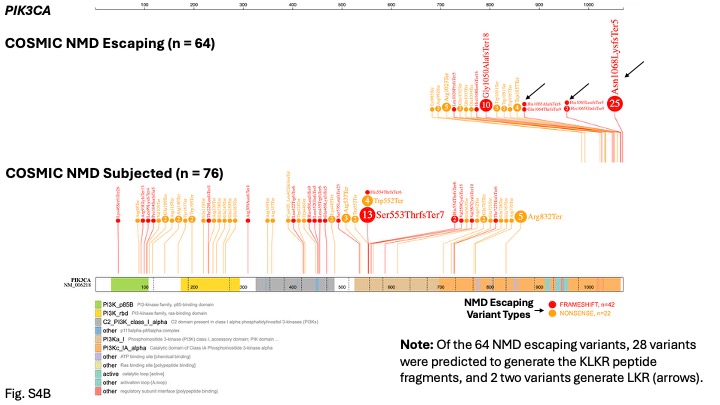

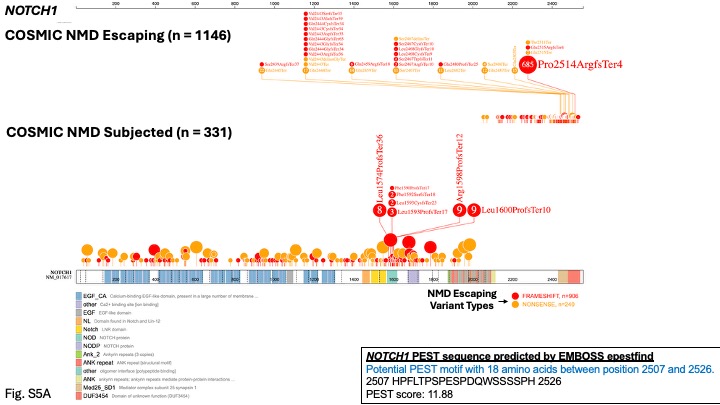

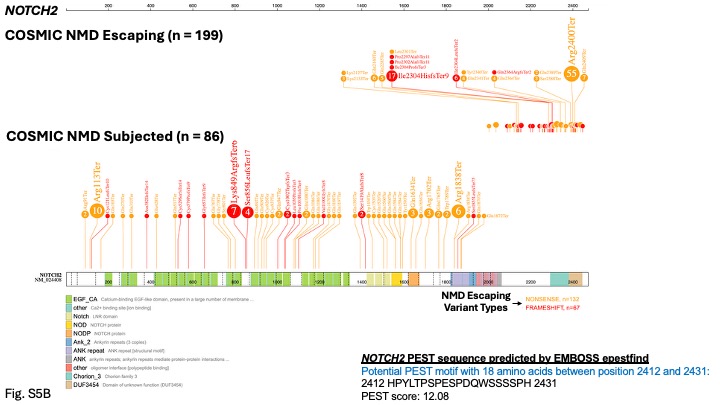

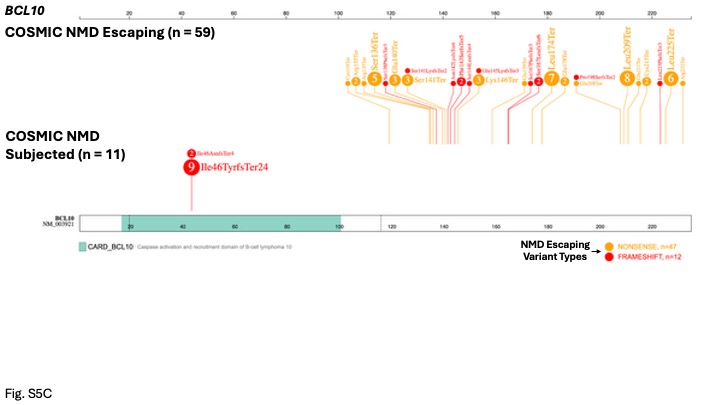

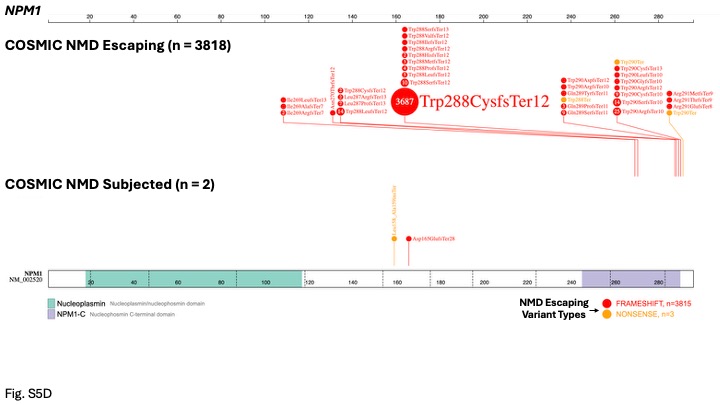

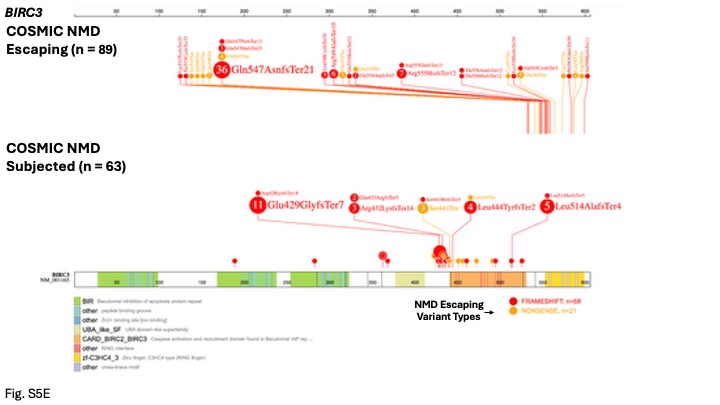

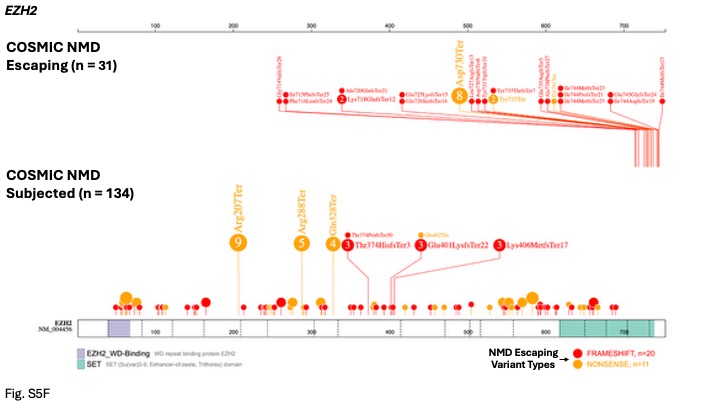

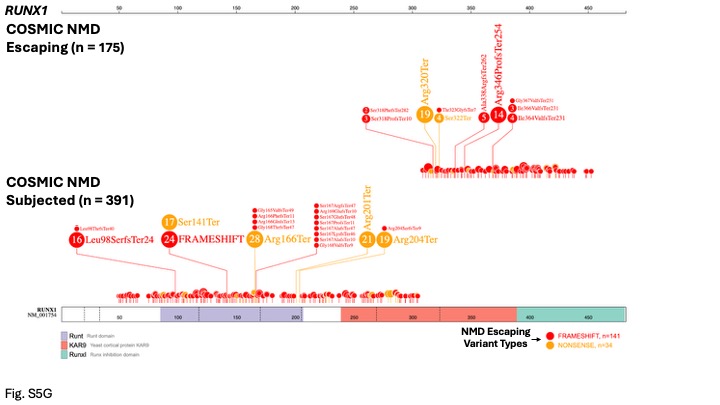

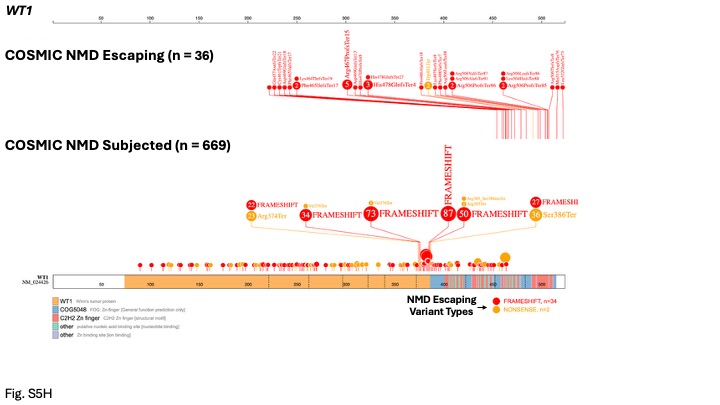

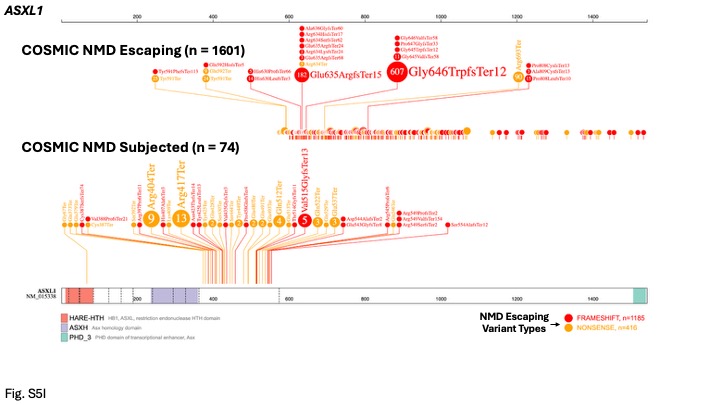

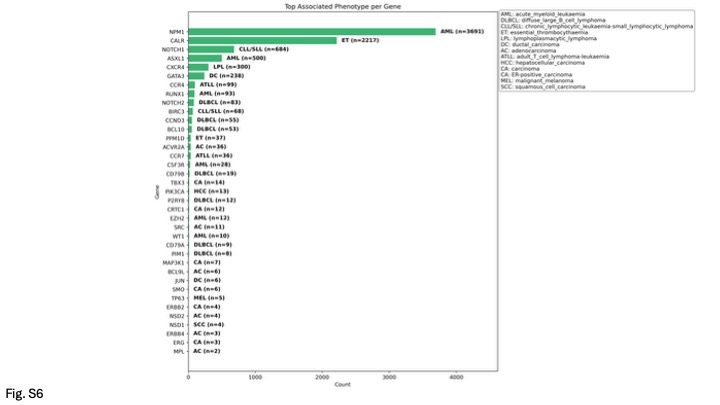

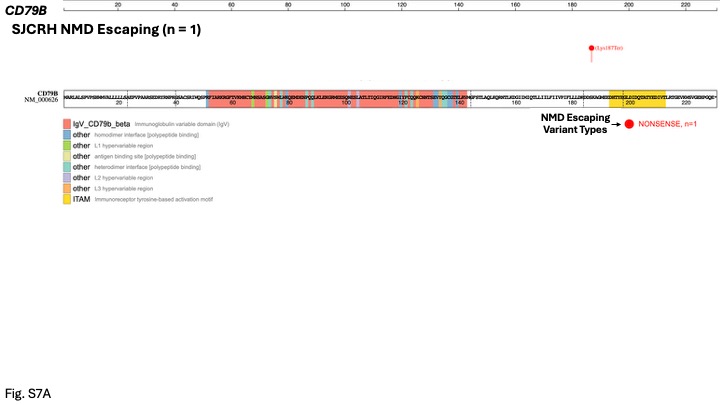

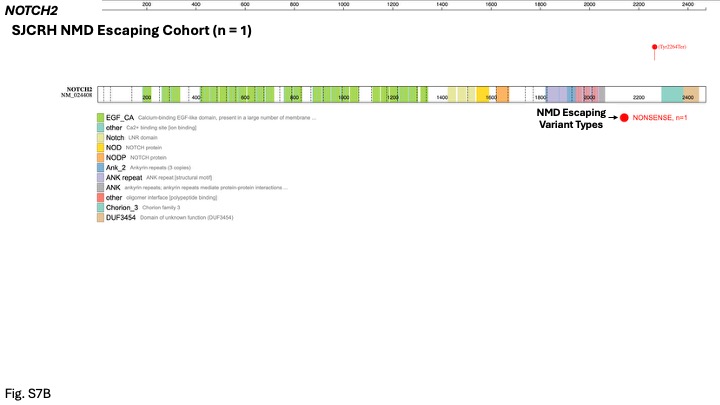
